# Assessing the plausibility of supercritical transmission for an emerging or re-emerging pathogen

**DOI:** 10.1101/2020.02.08.20021311

**Authors:** Seth Blumberg, Phoebe Lu, Thomas M Lietman, Travis C Porco

## Abstract

Rapid assessment of the transmission potential of an emerging or reemerging pathogen is a cornerstone of public health response. A simple approach is shown for using the number of disease introductions and secondary cases to determine whether the upper bound of the reproduction number exceeds the critical value of one.

## Objective

We present a simple analysis for monitoring the upper bound of estimates for the reproduction number, *R*, in settings where there are very few cases of disease. We focus particular attention on the threshold at which endemic transmission appears possible. This is relevant for deciding whether observed transmission events are sufficiently infrequent so that disease spread is selflimiting.

## Background

Subcritical transmission is characterized by *R* less than one. This signifies that on average, each new case causes less than one infection and so disease spread is self-limited. In contrast, supercritical transmission with *R* greater than one leads to the potential for endemic or epidemic transmission.

As the COVID-19 pandemic has illustrated, the classification of disease spread as being subcritical or supercritical has significant public health implications for characterizing the risk of emerging infections.^1^ When control interventions are enacted for a disease with supercritical transmission, identifying if and when transmission becomes subcritical is an important indicator of the effectiveness of public health interventions. In addition, this classification is a useful characterization of the risk of re-emergence of previously controlled disease that may be seen with novel strains of a pathogen or a change in vaccination effectiveness.

For diseases in which spread is on the cusp of the subcritical versus supercritical boundary, epidemiologic investigations typically include an assessment of whether a new case is a ‘primary case’ due to introduction of disease into a population of interest, or a ‘secondary case’ due to spread of disease within that population.^2^ The number of disease introductions and subsequent transmissions can be utilized to infer a range for the reproduction number. If the upper confidence interval of this range is below one, subcritical transmission is likely.

## Methods and Findings

Our method draws from prior work that showed that the number of cases caused by each infection is effectively modelled via a negative binomial offspring distribution.^3^ This approach incorporates both the strength and heterogeneity of disease transmission. With this model, the maximum likelihood estimation of the reproduction number, *R*, is simply the proportion of cases due to community transmission (Figure 1A).^2,4^ The upper bound of the confidence interval for the *R* estimate can also be determined with this minimal data, provided that an assumption is made for the amount of heterogeneity in disease transmission, as quantified by the dispersion parameter, *k* (Figure 1B). The smaller *k* is, the more transmission heterogeneity there is and the larger the upper bound of the confidence interval. To minimize the possibility of underestimating the risk of supercritical transmission, we use *k* = 0.2 as our default value, consistent with the lower values of dispersion seen in the literature.^3^ As the number of observed cases increases for a fixed proportion of secondary cases, the maximum likelihood estimate of *R* remains constant. Because the precision of inference improves, the upper bound of the confidence interval decreases. For example, if there are 5 observed transmissions with 10 introductions or 10 observed transmissions with 20 introductions, the maximum likelihood estimate of *R* is 0.33. However, the upper bound of the 95% confidence interval has a supercritical value 1.62 in the former scenario and a subcritical value of 0.98 in the latter scenario.

**Figure 1:**
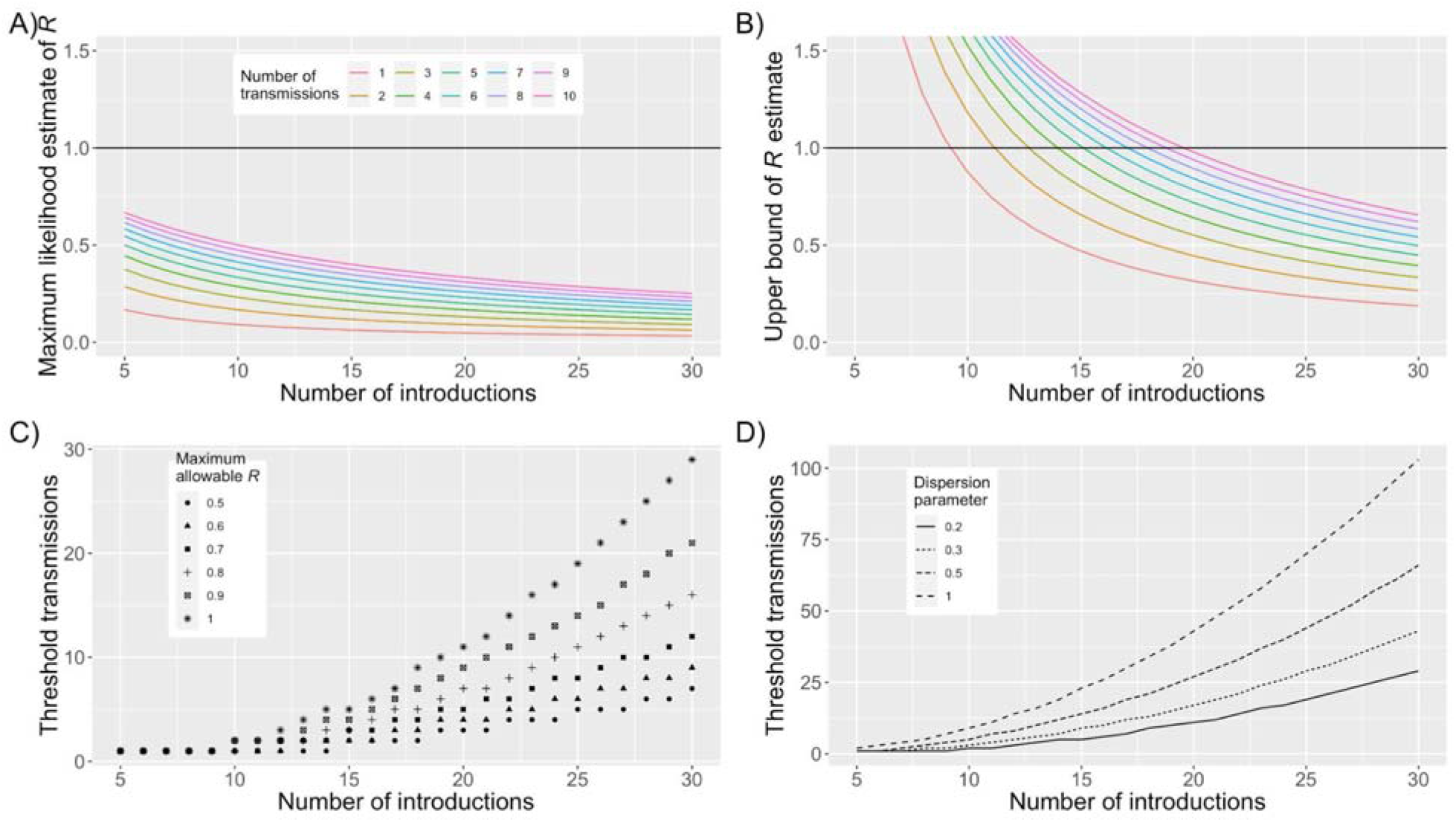
Evaluating the plausibility of subcritical transmission. A) The maximum likelihood estimate for the reproduction number, *R*, as a function of the number of disease introductions and linked transmissions. The black line corresponds to the critical value of *R* = 1 that distinguishes self-limited, subcritical transmission from sustained, supercritical transmission. These particular results are independent of the dispersion parameter which quantifies the degree of transmission heterogeneity. B) The upper bound of the 95% confidence interval for the reproduction number as a function of the number of disease introductions and linked transmissions. The colors correspond to the same legend as in panel A. A dispersion parameter of 0.2 is assumed, which corresponds to a high possibility of superspreading activity. C) The maximum number of transmissions that can be observed without concern that *R* has exceeded a predetermined threshold. The legend reflects the predetermined threshold, by indicating the maximum allowable value for the upper bound of the 95% confidence interval for *R*. A dispersion parameter of 0.2 is assumed. D) The number of secondary cases that can be observed before endemic transmission is possible (i.e. upper bound of 95% confidence of *R* is greater than 1). The plot shows the dependence on several values of the dispersion parameter as indicated by the legend. The solid curve corresponds to the results seen in panels B and C. Please see the main text and supplement for methodological details. An interactive display of results can be accessed at https://mindscape.shinyapps.io/Estimating_the_upper_limit_of_R/

From a public health perspective, it may be useful to model how many secondary cases can be observed before endemic transmission is statistically possible (Figure 1C). The threshold of secondary cases that can occur before endemic transmission is possible depends on the dispersion parameter, because higher values of the dispersion parameter narrow the confidence interval of *R* estimation (Figure 1D).^2^ In contrast to our default choice of 0.2 for the dispersion parameter which assumes superspreading is possible, many models are based on the homogeneous susceptible-infected-recovered framework that has a dispersion of one and thus predicts a much higher allowable number of transmission events before supercritical transmission is a concern. For example, when there are twenty disease introductions, the thresholds of secondary cases for supercritical transmission are 11 and 43 for dispersion values of 0.2 and 1, respectively. The value of our method in assessing the risk of endemic spread is illustrated for COVID-19 transmission in Hawaii in the early stages of the pandemic (Table 2).

**Table 1:**
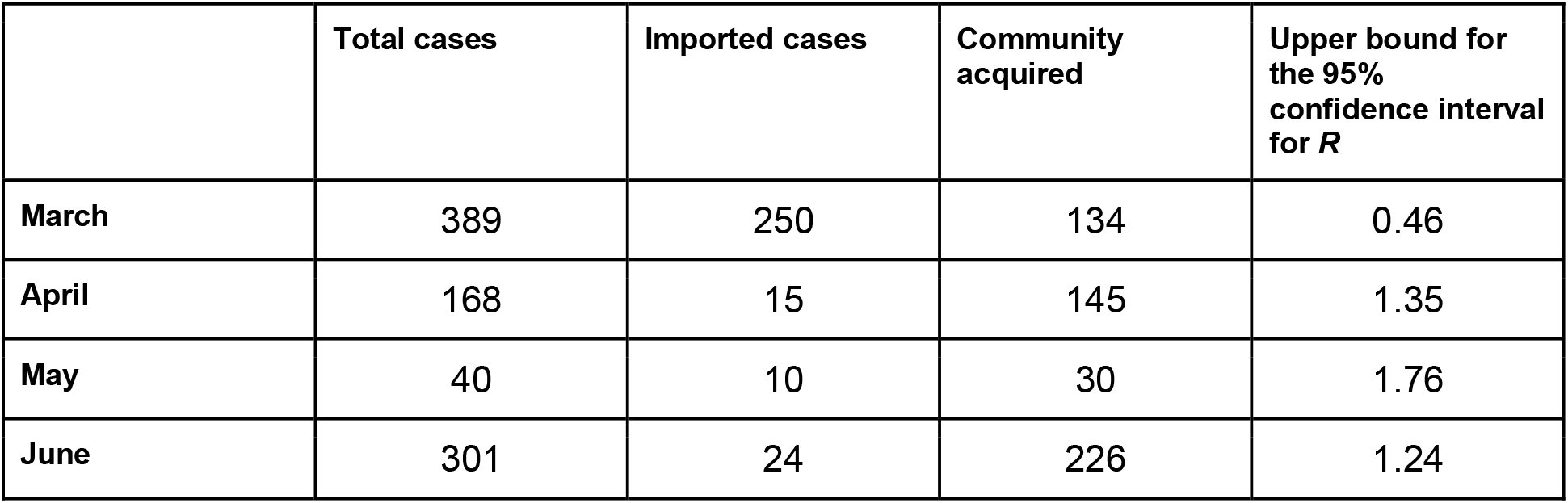
Assessing the potential for supercritical transmission in Hawaii. Early in the pandemic, when lockdowns were strictest, the potential for persistent disease spread appeared to be well controlled. Although the total number of cases decreased in April and May, the concern for supercritical spread increased. The relatively high proportion of cases due to community spread in May foreshadows the explosive increase in cases seen in June. Case counts were obtained from Hawaii’s Department of Health.^5^ The sum of imported and community-acquired cases does not always equal the total cases because some cases were categorized as having an unknown source.

| | Total cases | Imported cases | Community acquired | Upper bound for the 95% confidence interval for $R$ |
| --- | --- | --- | --- | --- |
| March | 389 | 250 | 134 | 0.46 |
| April | 168 | 15 | 145 | 1.35 |
| May | 40 | 10 | 30 | 1.76 |
| June | 301 | 24 | 226 | 1.24 |

**Table 2:** Evaluating the impact of imperfect observation. Statistics for the distributions seen in Figure 2. The median values for each distribution are shown. Numbers in parentheses are the 2.5th and 97.5th quantiles for the distributions.

| Observation scenario | Passive : active observation probabilities | Number of observed transmissions | Maximum likelihood value of $R$ | Upper bound for the 95% confidence interval for $R$ | Fraction of simulations where supercritical transmission appears plausible |
| --- | --- | --- | --- | --- | --- |
| Passive observation only | 0.5 : 0.0 | 19 (4 - 52) | 0.39<br>(0.12 - 0.63) | 0.87<br>(0.39 - 1.14) | 25% |
| Partial contact tracing | 0.5 : 0.5 | 31 (8 - 82) | 0.51<br>(0.21 - 0.73) | 1.02<br>(0.58 - 1.20) | 57% |
| Perfect contact tracing | 0.5 : 1.0 | 43 (13-115) | 0.59<br>(0.30 - 0.79) | 1.10<br>(0.74 - 1.21) | 77% |
| Perfect observation | 1.0 : 1.0 | 25 (5 - 82) | 0.46<br>(0.14 - 0.73) | 0.96<br>(0.45 - 1.20) | 44% |

One caveat of our approach is that imperfect observation of cases can bias results.^4,6,7^ To obtain a sense of how imperfect observation affects results, we simulated observed transmission events for four observation scenarios and determined the corresponding results for inference of *R* (Figure 2, and Table 2). The scenarios are each represented by a passive and active observation probability.^2,6^ The passive observation probability reflects the chance that any case will be observed without enhanced surveillance. The active observation probability is the probability that a case will be observed only because of contact tracing resulting from passive observation of a linked case. The four scenarios we consider are passive observation only, partial contact tracing, perfect contact tracing, and perfect observation.

**Figure 2:**
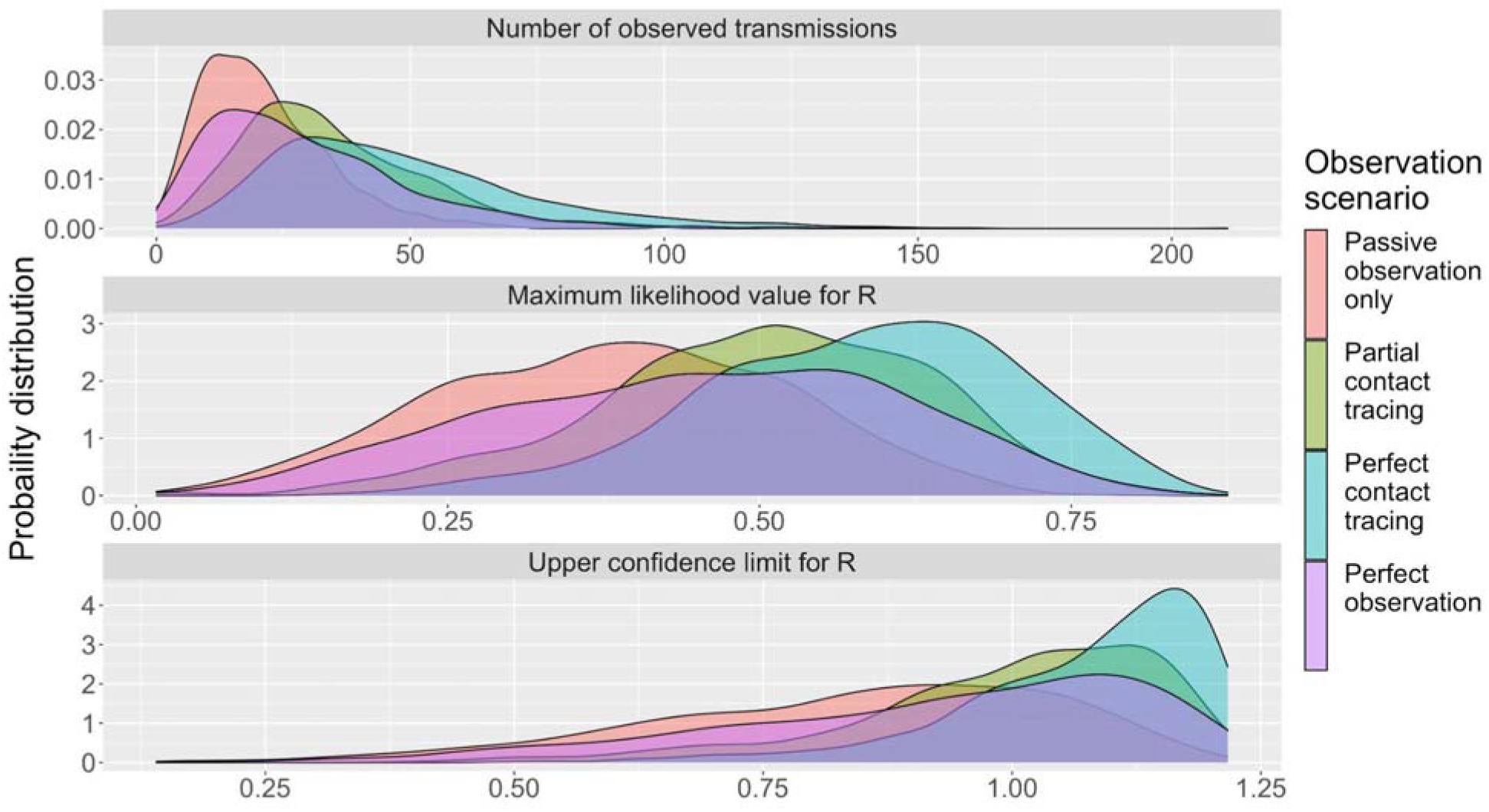
Evaluating the impact of imperfect observation. The top panel shows the probability distributions for the number of observed transmission events for different observation scenarios. The observation scenarios are determined by the passive and active observation probabilities, as indicated by Table 2. Each histogram represents results when thirty observed transmission chains are simulated. A reproduction number of 0.5 and a dispersion parameter of 0.2 is assumed. Results are based on 1,000 simulations. The middle panel shows the distributions for the maximum likelihood estimate of *R*, based on the simulations generated for the top panel. The bottom panel shows the corresponding distributions for the upper value of the 95% confidence interval for the inferred value of *R*. Please see the main text and supplement for methodological details.

We find that when compared to perfect observation, passive observation decreases the observed number of transmissions and leads to an underestimation of the risk of supercritical transmission. In contrast, imperfect observation with perfect contact tracing of passively observed cases has the opposite effect. This is because passively observed cases are more likely to be in large transmission chains and thus contact tracing biases the observed transmission chain size upwards.^2,6^ Thus, if contact tracing is robust then the bias introduced will overestimate the plausibility of subcritical transmission.

## Discussion

While the reproduction number is a useful indicator for the transmissibility of a disease, it often varies amongst subpopulations and over time. Variation in climate, population density, demographics, social interactions, health care access, and public health interventions can all affect transmission. The approach we have presented for evaluating if the observed number of disease introductions and transmission events is consistent with supercritical transmission is particularly applicable for monitoring disease in well-defined populations such as geographically-restricted regions, hospitals, prisons, or dormitories. The COVID-19 pandemic has been the main driver for development of this approach, as public health agencies wrestle with knowing when local control is achieved for specific communities and working environments. However, this methodological approach is also applicable to other diseases exhibiting human-to-human spread.

Importantly, this approach assumes that none of the observed cases remain infectious and that there are no patients who are already infected but are pre-symptomatic. This may be a reasonable assumption if transmission primarily occurs when patients are symptomatic and if symptomatic patients are quickly quarantined. However, until there has been a substantial gap in time (at least one serial interval) since a patient is quarantined, this assumption requires judicious consideration.

## Supporting information

Supplement: Description of methods

## Data Availability

This manuscript is focused on analysis of publicly available data.

https://mindscape.shinyapps.io/Estimating_the_upper_limit_of_R/

https://github.com/proctor-ucsf/supercritical_plausibility

## Funding

SB and TCP are supported by CDC U01CK000590, as part of the Modeling Infectious Diseases in Healthcare Network.

## Notes

### Competing Interest Statement

The authors have declared no competing interest.

### Funding Statement

None of the authors received payment or services from a third party for any aspect of the submitted work. The authors are supported by grants by the NIH and Gates Foundation, but no additional funding was received for this work.

### Summary of Updates

Key updates: 1. Context of methodological approach is clarified 2. Specific example of COVID spread in Hawaii is highlighted 3. Sensitivity analysis added for imperfect case observation

