## Supplement: Description of methods for "Assessing the plausibility of supercritical transmission for an emerging or re-emerging pathogen"

November 3, 2021

All calculations and simulations are conducted in R version 4.1.1. The code is provided at

https://github.com/proctor-ucsf/supercritical_plausibility.

### 1 Inference of R

Based on prior research, we assume that the every new infection causes subsequent infection according to a negative binomial distribution with mean, $R$ and dispersion parameter $k$.[1] With this assumption, the likelihood that $i$ cases cause $j$ cases in a single generation of transmission is,[2,3]

$$L_{i\to j} = \frac{\Gamma(j+ki)}{\Gamma(j+1)\Gamma(ki)}\left(\frac{k}{R+k}\right)^{ki}\left(\frac{R}{R+k}\right)^{j}. \tag{1}$$

The likelihood that $m$ primaries causes $n$ total cases over multiple generations is then,[2,4,5]

$$L^{C}_{m\to n} = \frac{m}{n}L_{n\to(n-m)}. \tag{2}$$

Maximizing the likelihood reveals that the maximum likelihood value of $R$ is $\frac{n-m}{n}$, which is simply the proportion of cases due to secondary transmission (i.e. non-imported cases).[6] The confidence interval for $R$ is obtained by the likelihood ratio test, which models the log of the likelihood function as following a chi-squared distribution.[7] The threshold number of transmissions indicating concern that $R$ may have exceeded a specific value (Figure 1C and 1D of the main text) was determined by incrementally increasing the number of observed transmissions and observing when the inferred confidence interval for $R$ crossed the threshold value.

### 2 Notable caveats

Notable caveats to these equations are that the $R$ and $k$ do not change from generation to generation, and that all cases are observed. Processes such as depletion of susceptible individuals or isolation can decrease $R$ changes in late generations of a transmission chain. Such changes would lead to an overestimation of the capacity for sustained transmission, and in this context our estimates for the probability of sustained transmission are conservative.

Imperfect case observation has more complicated implications for the bias of our $R$ estimates. A particular concern is if, at the time of data collection, many cases remain infectious. This would imply that the full potential full endemic transmission may not yet have been demonstrated and could lead to underestimation of the capacity for sustained transmission. A second concern is that cases do not always reach the attention of surveillance systems. We address this possibility in the next section.

### 3 Simulation of imperfect observation

We model two possible ways that cases may be observed. The first is that there is an independent probability of $p_{passive}$ that any given case will be observed by the surveillance system. Once one or more cases in a chain is observed by this 'passive' process, we assume the other cases in the transmission chain can be observed via an 'active' contact tracing process with an independent probability of $p_{active}$.[6,8] Perfect observation corresponds to $p_{passive} = 1$. Absence of contact tracing corresponds to $p_{active} = 0$, and perfect contact tracing corresponds to $p_{active} = 1$.

To conduct our analysis of imperfect observation, we run 1,000 simulations of observation scenarios corresponding to specific values of $p_{passive}$ and $p_{active}$. The output of each simulation is an observed number of secondary transmission events for a pre-specified number of observed importations. For each simulation run, we infer the corresponding confidence interval for $R$. We use the distribution of these inferred confidence intervals to examine the impact of imperfect observation via Table 2 of the main text.
